# Changes in Reproductive Rate of SARS-CoV-2 Due to Non-pharmaceutical Interventions in 1,417 U.S. Counties

**DOI:** 10.1101/2020.05.31.20118687

**Authors:** Jie Ying Wu, Benjamin D. Killeen, Philipp Nikutta, Mareike Thies, Anna Zapaishchykova, Shreya Chakraborty, Mathias Unberath

## Abstract

In response to the rapid spread of the novel coronavirus, SARS-CoV-2, the U.S. has largely delegated implementation of non-pharmaceutical interventions (NPIs) to local governments on the state and county level. This staggered implementation combined with the heterogeneity of the U.S. complicates quantification the effect of NPIs on the reproductive rate of SARS-CoV-2.

We describe a data-driven approach to quantify the effect of NPIs that relies on county-level similarities to specialize a Bayesian mechanistic model based on observed fatalities. Using this approach, we estimate change in reproductive rate, *R_t_*, due to implementation of NPIs in 1,417 U.S. counties.

We estimate that as of May 28^th^, 2020 1,177 out of the considered 1,417 U.S. counties have reduced the reproductive rate of SARS-CoV-2 to below 1.0. The estimated effect of any individual NPI, however, is different across counties. Stay-at-home orders were estimated as the only effective NPI in metropolitan and urban counties, while advisory NPIs were estimated to be effective in more rural counties. The expected level of infection predicted by the model ranges from 0 to 28.7% and is far from herd immunity even in counties with advanced spread.

Our results suggest that local conditions are pertinent to containment and re-opening decisions.

## 1 Introduction

As of May 28^th^, 2020, the United States has reported more than 1,700,000 cases of novel coronavirus 2019 (COVID-19). This disease, caused by severe acute respiratory syndrome coronavirus 2 (SARS-CoV-2) infection, has led to more than 100,000 deaths in the U.S.^1^ Non-pharmaceutical interventions (NPIs) are a critical component of the public health effort to slow the spread of COVID-19 as pharmaceutical interventions are not currently available. NPIs include guidelines for hand hygiene, cancellations of mass events, school closures, closure of non-essential business, and stay-at-home orders. These measures are designed to reduce transmission, buy time to expand healthcare capacity, develop effective testing and tracing mechanisms, and research pharmaceutical options, such as a vaccine.

Indeed, the implementation of NPIs caused a rapid decline of new cases and deaths, although at a very high economic and social cost. Understanding precisely how NPIs affect disease transmission is desirable in order to safely roll back NPIs and minimize adverse effects. Prior works have quantified the effects of NPIs on the reproductive rate *R_t_* of SARS-CoV-2 in China,^2^ UK,^3^ Brazil,^4^ and 14 European countries, including Italy, Spain, and Germany.^5,6^ Except for some large urban areas,^7^ these effects have not yet been quantified for the U.S., which delegated NPI implementation to state and local governments rather than establishing a unified, federal approach. As a result, it is necessary to quantify the effect of NPIs on the county level, but the limited number of documented fatalities in many counties would limit this analysis to urban areas with sufficient data for epidemiological modeling.

We develop a data-driven approach that establishes county similarity based on factors known to affect community transmission of infectious diseases. Within groups of similar counties, we jointly optimize the parameters of a Bayesian mechanistic model to the observed deaths in every county under the assumption that the same NPI attains comparable effects across counties that are similar with respect to disease transmission factors. Using this approach, we estimate the change in *R_t_* due to implementing NPIs in 1,417 U.S. counties that exhibit substantially different characteristics regarding population density, economy, demographics, and infrastructure. We fit a model to groups of counties with similar conditions which gives us a probable *R_t_* trajectory over time and consequently, the probable number of people infected over time.

## 2 Conclusions

Based on the model, as of May 28^th^, 2020 1,177 out of the considered 1,417 U.S. counties are estimated to have reduced the reproductive rate of novel coronavirus to below 1.0 via the implementation of NPIs. The estimated effect of any individual NPI, however, may differ across counties. We observe that for metropolitan and urban counties, the most substantial reduction is attributed to stay-at-home orders while less restrictive NPIs were estimated to be effective in more rural counties. Further, the expected level of infection predicted by the model, ranging from 0 to 28.7%, is far from herd immunity even in counties with advanced spread.

While the model explains the observed trends in fatalities well, the rapid escalation of the COVID-19 situation and quick succession in the implementation of NPIs in most counties within few days from another complicates the disentanglement of the effects of any individual NPI. Further, even if accurate, the effect attributed to any NPI may not describe the consequences of roll back of the same NPI because social norms and individuals’ behavior may have changed, including the wearing of masks in public.

Despite these limitations, our results suggest that strategies for shutdown as well as re-opening require careful consideration of county conditions in addition to state and national trends.

## 3 Results

### Characterizing Groups of Similar Counties as Clusters

Since our hypothesis is that local conditions affect the spread of COVID-19, we differentiate among groups of counties using a data-driven approach known as clustering. Each group – or cluster – of counties is characterized by having similar demographic and socioeconomic qualities. For instance, cluster 1 consists of low-population, mostly rural counties with little public transit capacity and the lowest median household income. Cluster 2 and 3 are similar in size, having a mean population of 45,000 and 52,000 respectively, but cluster 2 includes higher-income, suburban areas where-as cluster 3 has lower income areas with a large land area. Cluster 4 consists of the densest metropolitan areas with a high proportion of 18- to 65-year-olds, high household income, a high public transit score, and small land area. Finally, cluster 5 has the highest mean population besides cluster 4, but is less densely populated, has poor public transit, and a lower household income on average. It is important to note that although we refer to these clusters with numbers 1–5, these are merely labels returned by the clustering algorithm without any meaning inherent to the ordering Figure 1. shows our clustering for all U.S. counties with this data available, including those without any incidence of COVID-19.

**Figure 1:**
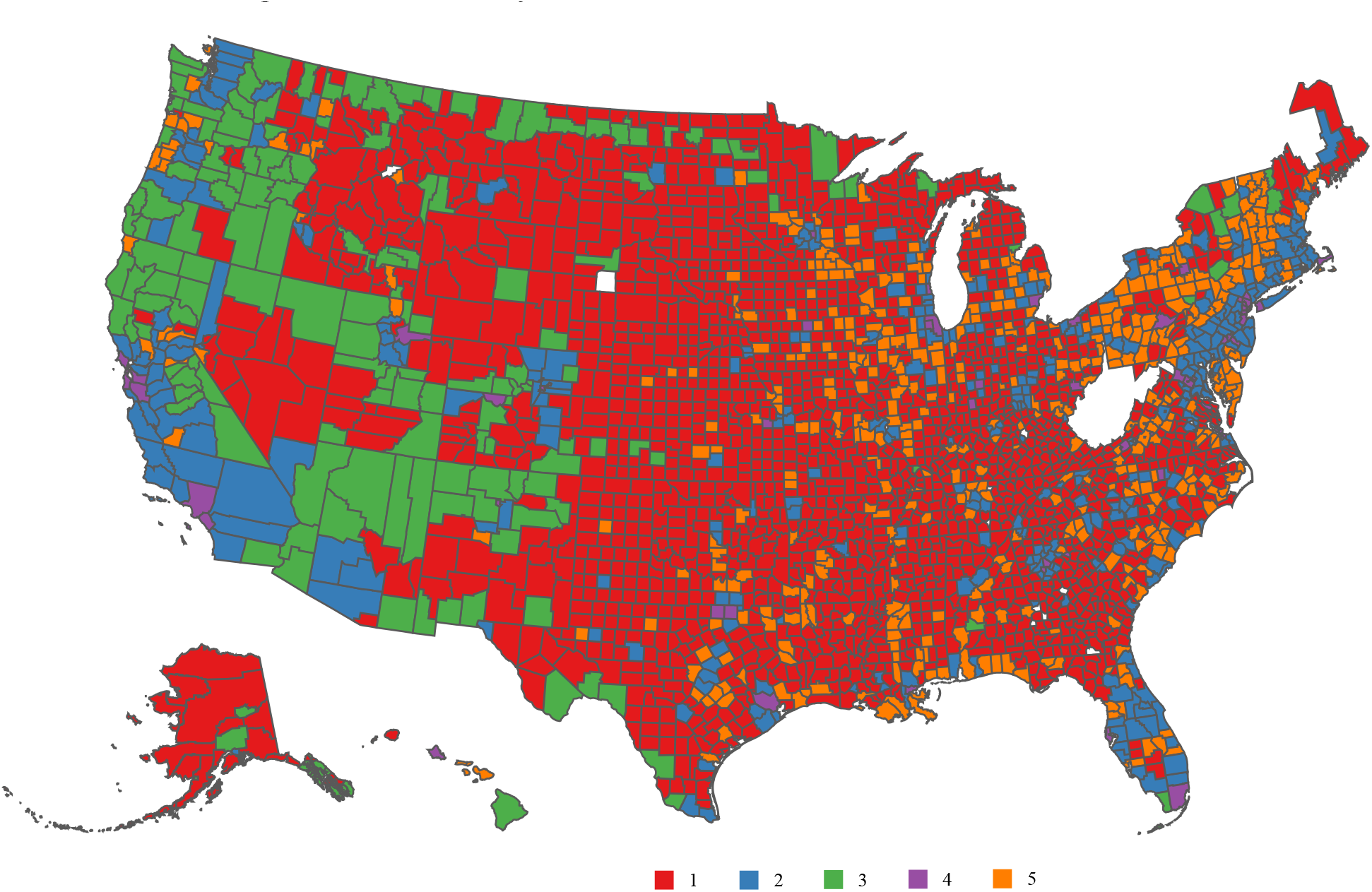
Cluster labels based on demographic and socioeconomic conditions are used to aggregate data and specialize epidemiological models. Here, one can see how cluster 1 and 3 primarily cover rural areas, while clusters 5, 2, and 4 consist of increasingly urban counties.

Once we have identified the clusters, we infer the reproductive rate of SARS-CoV-2 over time. by jointly optimizing the parameters of a Bayesian mechanistic model on each cluster. This process is described in greater detail in Section 4. Figure 2 shows an example of this progression for counties ordered by their public transportation use.

**Figure 2:**
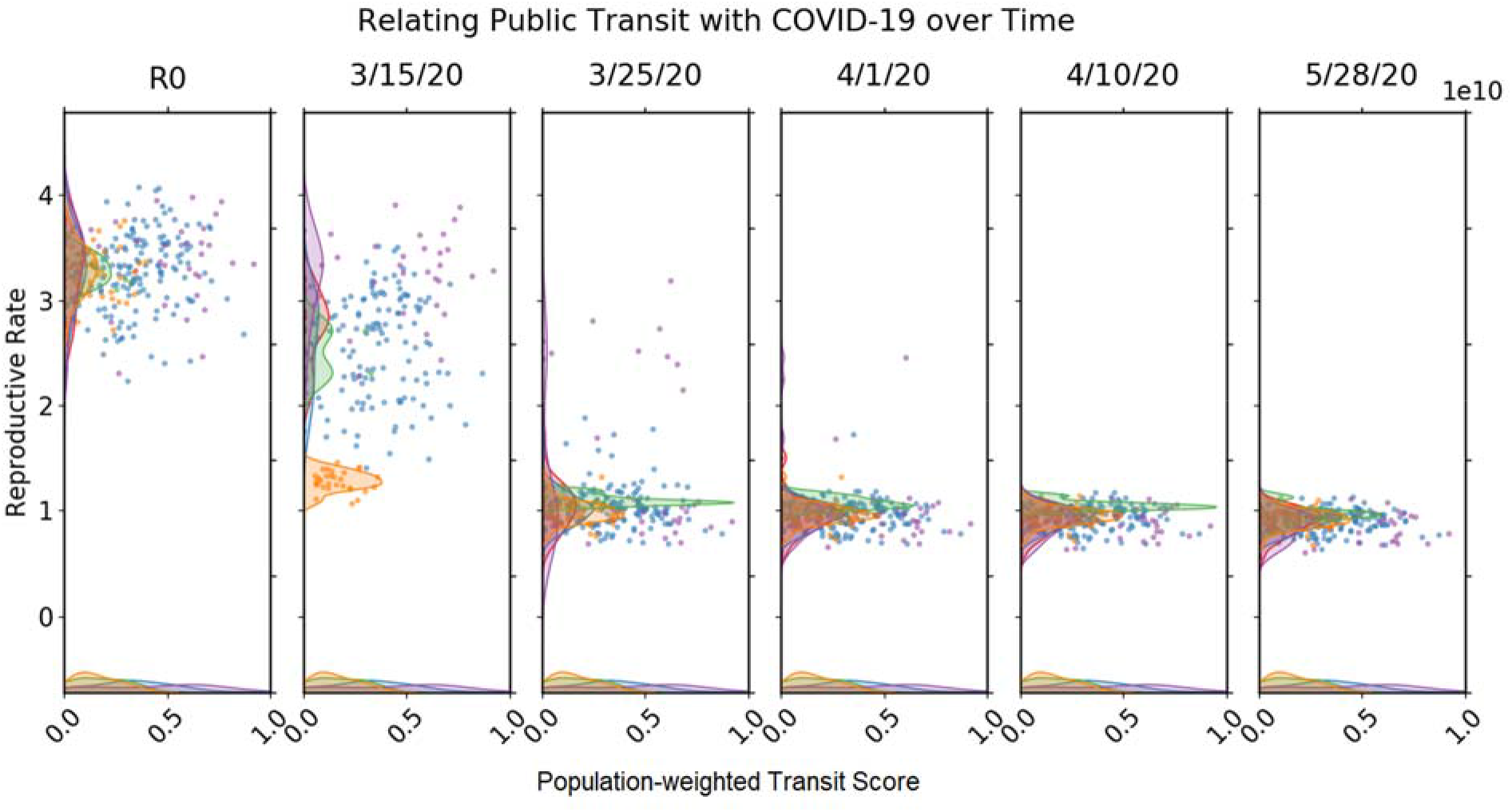
Relationship between public transit capacity and the time-dependent reproductive rate of SARS-CoV-2 for U.S. counties. Colors correspond to clusters as in Figure 1. In the cluster of counties that rely least on public transport, shown in yellow, we observe a substantial drop in reproductive rate already on 3/15/20, possibly indicating a strong effect of advisory NPIs that were in effect at that time.

We observe that although the basic reproductive rate of SARS-CoV-2 starts at a similar level for all clusters, the speed at which counties in each cluster respond to the disease, and reduce its *R_t_* differs. The reproductive rate in metropolitan counties (cluster 4), which tends to have higher reliance on public transportation decreases over the entire period. This is especially apparent going from March 15 to March 25. On the other hand, clusters with little transportation use, such as cluster 5, was estimated to have quickly reduced transmission rates. This suggests that if people have a higher reliance on public infrastructure, more stringent interventions are necessary to reduce transmission. The staggered drops in reproductive rates are attributed to both varied efficacy of NPIs (to be discussed in the next section) and counties implementing NPIs at different times. Comparisons with more features are shown in Section 4.

### Estimates of Initial and Current Reproductive Rates and Number of Infected

From Table 1, we observe that most counties exhibited an initial reproductive rate *R*_0_ above 3. As of May 28^th^, however, most have successfully reduced the reproductive rate to *R_t_* ≈ 1 after implementing NPIs. It is estimated that 18.4 to 42.7% of the population (95% confidence interval) has been infected in New York, NY, which has the highest number of cases. Based on the initial reproductive rates between 2 and 4, herd immunity is reached only after 50 – 70% of the population has recovered,^8,9^ suggesting that all U. S. counties are far from resilience. Consequently, easing restrictions is likely to result in subsequent waves of the epidemic. We state these findings for 15 heterogeneous counties in Table 1 and provide the same metrics for all 1,417 counties online at https://github.com/JieYingWu/npi-model, where we also provide code and data required for reproduction.

**Table 1:** Estimated initial and current reproductive rate as of May 28^th^, 2020, and the number of cases for select counties ordered by their reproductive rate as of May 28^th^. This is compared to the measured number of cases and fatality rates.

| County | Cluster | $R_0$<br>(std) | $R_{5/28}$<br>(std) | # (%) infected as<br>predicted | Measured<br>cases | Fatality rate<br>(measured<br>death/cases) |
| --- | --- | --- | --- | --- | --- | --- |
| 36061<br>New York, NY | 4 | 2.95<br>(0.16) | 0.59<br>(0.03) | 2388875 (28.4) | 201051 | 10.65% |
| 11001<br>DC | 4 | 3.11<br>(0.21) | 0.78<br>(0.05) | 78504 (11.1) | 8492 | 5.33% |
| 25013<br>Hampden, MA | 2 | 3.29<br>(0.17) | 0.81<br>(0.04) | 72967 (15.5) | 5878 | 9.56% |
| 35031<br>McKinley, NM | 3 | 3.44<br>(0.29) | 0.81<br>(0.05) | 20722 (28.7) | 2291 | 4.36% |
| 09007<br>Middlesex, CT | 2 | 3.16<br>(0.23) | 0.82<br>(0.06) | 17266 (10.6) | 1082 | 13.31% |
| 42079<br>Luzerne, PA | 2 | 3.11<br>(0.23) | 0.85<br>(0.06) | 18675 (5.9) | 2689 | 5.17% |
| 22015<br>Bossier, LA | 5 | 3.17<br>(0.15) | 0.88<br>(0.04) | 3771 (3.0) | 406 | 6.40% |
| 13241<br>Rabun, GA | 1 | 3.34<br>(0.15) | 0.91<br>(0.03) | 138 (0.8) | 19 | 5.26% |
| 04005<br>Coconino, AZ | 3 | 3.20<br>(0.19) | 0.93<br>(0.04) | 17516 (12.3) | 1078 | 7.33% |
| 05029<br>Conway, AR | 1 | 3.30<br>(0.25) | 0.94<br>(0.06) | 141 (0.7) | 14 | 7.14% |
| 53077<br>Yakima, WA | 3 | 3.17<br>(0.19) | 0.97<br>(0.05) | 17829 (7.1) | 3231 | 2.88% |
| 06037<br>Los Angeles, CA | 4 | 3.68<br>(0.18) | 1.00<br>(0.04) | 401338 (4.0) | 49860 | 4.49% |
| 28157<br>Wilkinson, MS | 1 | 3.72<br>(0.19) | 1.01<br>(0.05) | 1461 (16.7) | 85 | 10.59% |
| 37125<br>Moore, NC | 5 | 3.44<br>(0.17) | 0.99<br>(0.05) | 1424 (1.4) | 220 | 4.55% |
| 48291<br>Liberty, TX | 5 | 3.58<br>(0.17) | 1.03<br>(0.05) | 570.45 (0.7) | 81 | 3.79% |

### Learned Effects of NPIs

Using a cluster-specialized model, we quantify the effectiveness of NPIs, as shown below in *Table 2* and Table 3.

**Table 2:** The learned weights of the interventions for each cluster-specialized model, showing mean with standard deviation in parentheses. Note how the interventions have different effects for the different clusters.

| *Intervention* | *Cluster 1* | *Cluster 2* | *Cluster 3* | *Cluster 4* | *Cluster 5* |
| --- | --- | --- | --- | --- | --- |
| *I1: Stay at home* | 0.018 (0.040) | 0.141 (0.095) | 0.036 (0.098) | **0.963 (0.165)** | 0.041 (0.061) |
| *I2: >50 gathering* | 0.192 (0.229) | 0.010 (0.037) | 0.096 (0.202) | 0.046 (0.089) | 0.057 (0.113) |
| *I3: >500 gathering* | 0.072 (0.124) | 0.332 (0.156) | 0.184 (0.286) | 0.020 (0.048) | 0.039 (0.085) |
| *I4: Public schools* | 0.081 (0.167) | 0.060 (0.137) | 0.112 (0.206) | 0.098 (0.149) | 0.038 (0.088) |
| *I5: Restaurant dine-in* | 0.068 (0.139) | 0.002 (0.024) | 0.148 (0.272) | 0.035 (0.082) | 0.021 (0.065) |
| *I6: Entertainment/gym* | 0.131 (0.175) | 0.005 (0.032) | 0.148 (0.255) | 0.033 (0.066) | 0.027 (0.066) |
| *I7: Federal guidelines* | **0.516 (0.394)** | **0.522 (0.283)** | 0.187 (0.305) | 0.060 (0.109) | 0.068 (0.147) |
| *I8: Foreign travel ban* | 0.181 (0.242) | 0.161 (0.182) | **0.192 (0.292)** | 0.011 (0.037) | **0.939 (0.200)** |

**Table 3:**
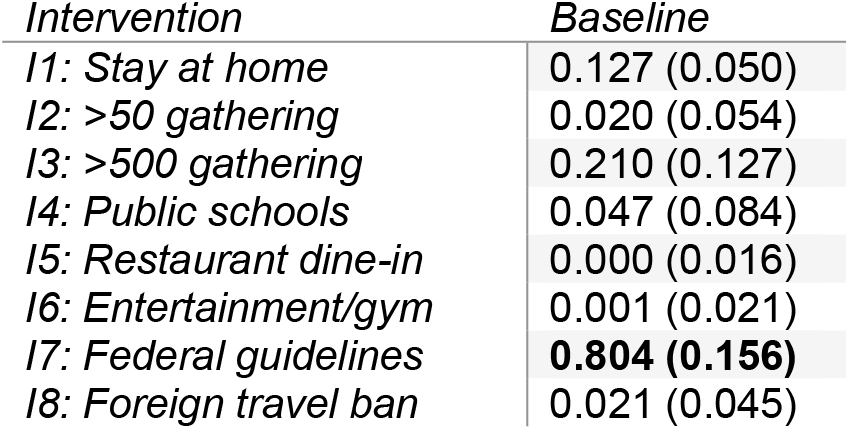
Without a cluster-specialized model, information about the effectiveness of local interventions is not as pronounced.

We note that our model estimates different behavior across counties. While all counties have implemented a similar set of interventions, their estimated effects are substantially different in each respective cluster. For example, metropolitan counties (cluster 4) were estimated to have a strong response to stay-at-home orders, while rural and suburban areas (clusters 1, 2, 3, and 5) responded more to national-level interventions according to our model.

One caveat is that the effects of interventions that were implemented close together are difficult to disentangle. Many local governments implemented formal NPIs in immediate response to the federal guidelines, leading a quick succession of NPIs coming into effect. Disentanglement of the effects of NPIs is explored more in Section 4.

Another limitation of our model is that the federal level interventions may have come before some counties have seen any cases. This introduces ambiguity between *R*_0_ estimates and the weight attributed to federal NPIs, i.e., federal guidelines and travel ban. Similar ambiguity is less prominent for other interventions, since they are not generally implemented before a county has seen cases. Additionally, since the epidemiological model used here is mechanistic, it only allows for changes of the transmission rate at the time of implementation of interventions. Other events, such as high-profile cases and cancellations of prominent festivals, likely contributed to increasing awareness of the disease and may also have effects on individual behavior, and therefore the *R_t_*. These effects cannot be attributed to a specific date and may thus affect the weights of interventions that come into effect at around the same time.

## 4 Methods

### Method Overview

Figure 3 shows an overview of the proposed method. We train a Bayesian mechanistic model (BMM) model in a cluster-specialized manner and compare with the baseline model trained on all counties. We show the stability of our model in two ways, first by withholding random days during training and second by withholding given counties. We compare the predictions based on incomplete data with those based on complete data.

**Figure 3:**
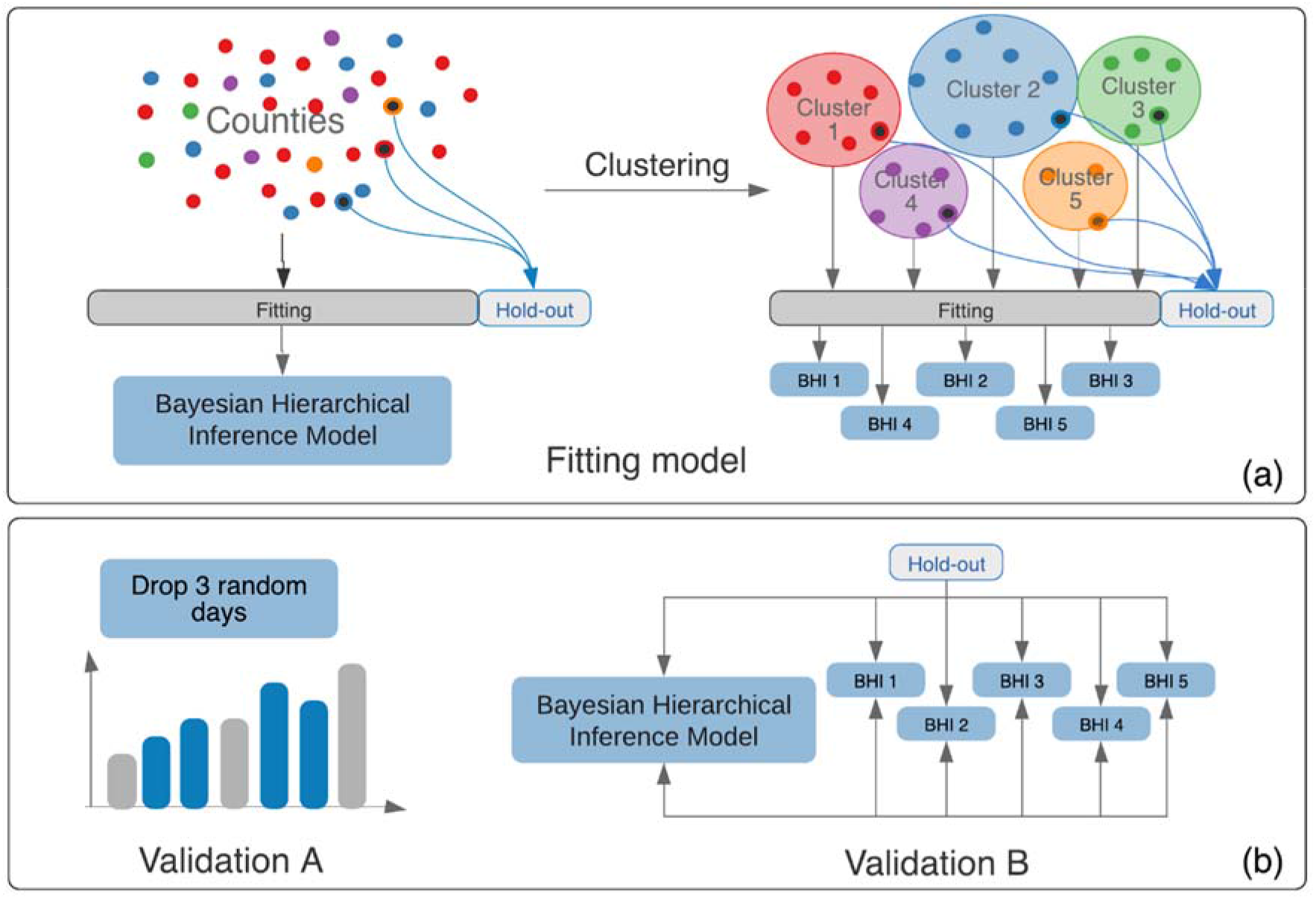
The fitting process for our model. We compare the performance of the baseline model **(a)**, which is fit to all eligible counties, with cluster-specialized models BHI 1–5. **(b)** BMM 1-5. **(c)** Two validation methods for our model. Validation A tests the stability of the model by withholding 3 random days and comparing the outputs. Validation B withholds a set of counties for comparison rather than days.

### County-level Clustering

Quantifying changes in COVID-19’s reproductive rate is complicated when considering differences at the county – rather than the national – level. Parametric epidemiological models are optimized to describe fatality counts, the volume and reliability of which decreases outside of highly populated regions. To make up for this scarcity, we leverage a balanced clustering of U.S. counties to aggregate data from similar counties in the same state, treating them as a single “super-county,” if they have identical NPI implementation dates. This has the advantage of considering counties that would otherwise be excluded without assuming that the spread of the disease in those counties follows the same trend of more advanced regions in the same state or country. Figure *4* shows how for a given state-in this case Texas, and a given cluster, the counties with 1–49 cumulative deaths from COVID-19 are treated as a single entity. In addition to this data aggregation strategy, we fit a cluster-specialized model to each set of counties, quantifying the possibly disparate effects of the NPIs in each type of county, as detailed below.

**Figure 4:**
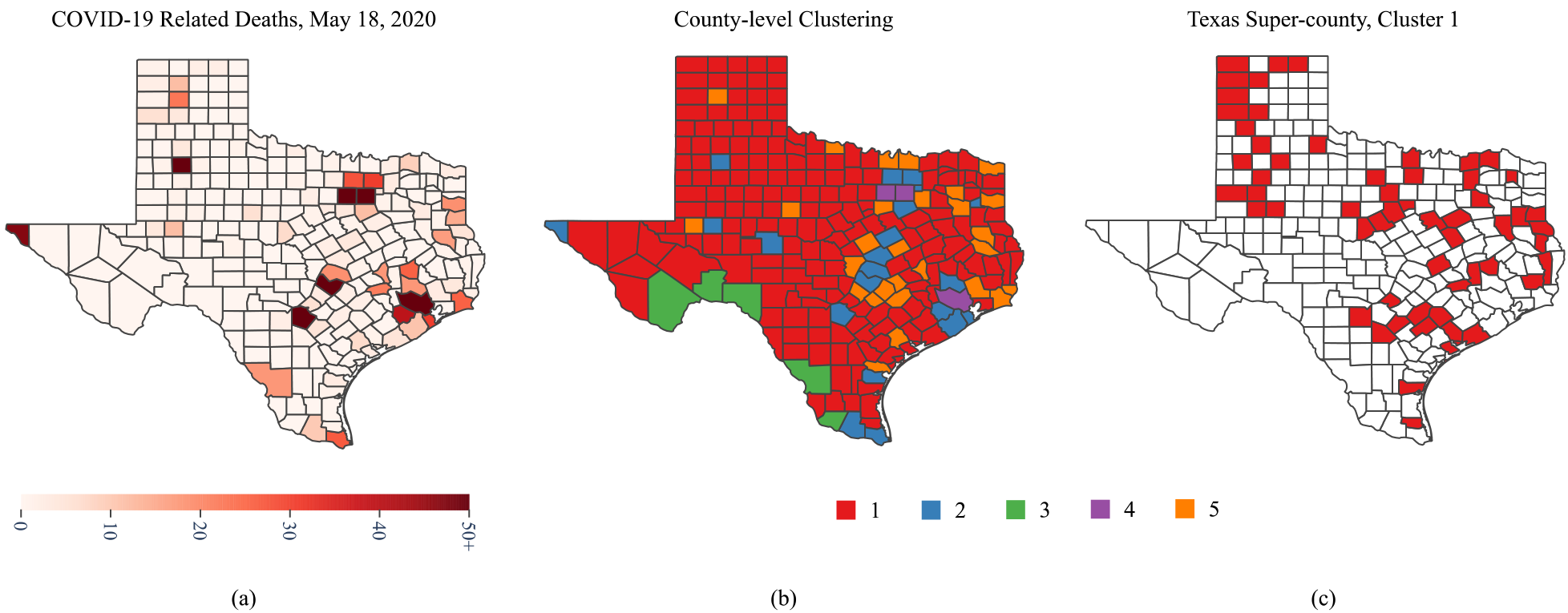
**(a)** The total confirmed deaths caused by COVID-19 for counties in Texas, as of May 28^th^. **(b)** Cluster labels for each Texas county, based on demographics, education, density, and other factors. **(c)** Texas counties in cluster 1 having 1–49 cumulative deaths as of May 28^th^, 2020 and the same NPI implementation dates.. To enable robust epidemiological models, these counties are treated as a single “super-county.”

To generate this clustering, we partition 3,059 U.S. counties into five clusters based on variables which directly affect disease spread, using a Gaussian mixture model.^10^ *Table 4* summarizes these variables, which include demographic, economic, and public transit capacities that we have gathered, processed for machine readability, and released in a publicly available dataset.^11^ Sources include the United States Census Bureau, the United States Department of Agriculture Economic Research Service and the Center for Neighborhood Technology. A full list of sources can be found on the corresponding website.^11^ To incorporate potential exposure, we consider county population density, housing density, and land area. Additionally, we consider portions of the population for age- and gender-based demographic categories, due to COVID-19’s disparate effects on these groups.^12–16^

**Table 4:** Average values for each of the 16 variables considered in our clustering, capturing demographic and socioeconomic information as well as transit and health care capacity.

| *Variable* | *Cluster 1*<br>*Avg. Value* | *Cluster 2*<br>*Avg. Value* | *Cluster 3*<br>*Avg. Value* | *Cluster 4*<br>*Avg. Value* | *Cluster 5*<br>*Avg. Value* |
| --- | --- | --- | --- | --- | --- |
| *Population* | 23,522.1 | 45,0789.2 | 52,425.3 | 759,468.1 | 84,594.5 |
| *Fraction of Population Male, age 0-17* | 0.114 | 0.116 | 0.113 | 0.105 | 0.110 |
| *Fraction of Population Female, age 0-17* | 0.108 | 0.111 | 0.108 | 0.101 | 0.105 |
| *Fraction of Population Male, age 18-64* | 0.297 | 0.304 | 0.298 | 0.320 | 0.300 |
| *Fraction of Population Female, age 18-64* | 0.2803 | 0.309 | 0.282 | 0.325 | 0.296 |
| *Fraction of Population Male, age 65+* | 0.093 | 0.071 | 0.097 | 0.064 | 0.085 |
| *Fraction of Population Female, age 65+* | 0.1075 | 0.088 | 0.102 | 0.085 | 0.103 |
| *Fraction of Population Number with Some*<br>*College or Associate's Degree* | 0.213 | 0.198 | 0.238 | 0.167 | 0.213 |
| *Fraction of Population in Poverty* | 0.153 | 0.109 | 0.146 | 0.140 | 0.135 |
| *Fraction of Population Unemployed* | 0.018 | 0.018 | 0.023 | 0.018 | 0.019 |
| *Median Household Income* | 49,085.13 | 69,118.17 | 53,606.34 | 68,624.46 | 5,4430.48 |
| *Population Density (persons per sq. mile)* | 42.3 | 626.6 | 18.9 | 3789.1 | 132.6 |
| *Number of Housing Units (per capita)* | 0.498 | 0.389 | 0.539 | 0.415 | 0.454 |
| *Land Area (sq. miles)* | 1,120.42 | 993.03 | 3,409.26 | 355.80 | 650.96 |
| *Population-weighted Transit Score* | 0 | 2.70e+07 | 1.18e+07 | 2.86e+07 | 1.13e+07 |

Our clustering is also based on socioeconomic variables, which may indicate behavioral traits relevant to the spread of COVID-19. For instance, workers with tertiary education are more likely to hold office-type jobs which can be done from home.^17^ At the same time, many secondary-education jobs have been deemed essential, resulting in a high contact rate, which in turn increases the likelihood of infection. Thus, our clustering considers college education, poverty, unemployment, and median household income for each county as a reflection of the overall job composition in the local area. Finally, we include a population-weighted transit score due to the likelihood of transmission in the enclosed, possibly crowded space that public transport entails.

Figure *5* and Figure *6* show the plotted over median income and density for counties and super-counties in the different clusters. Super-counties are visualized as a single point using their population-weighted average for that feature. The plots show the distribution of the cluster over the features and its correlation with how *R_t_* changes.

**Figure 5:**
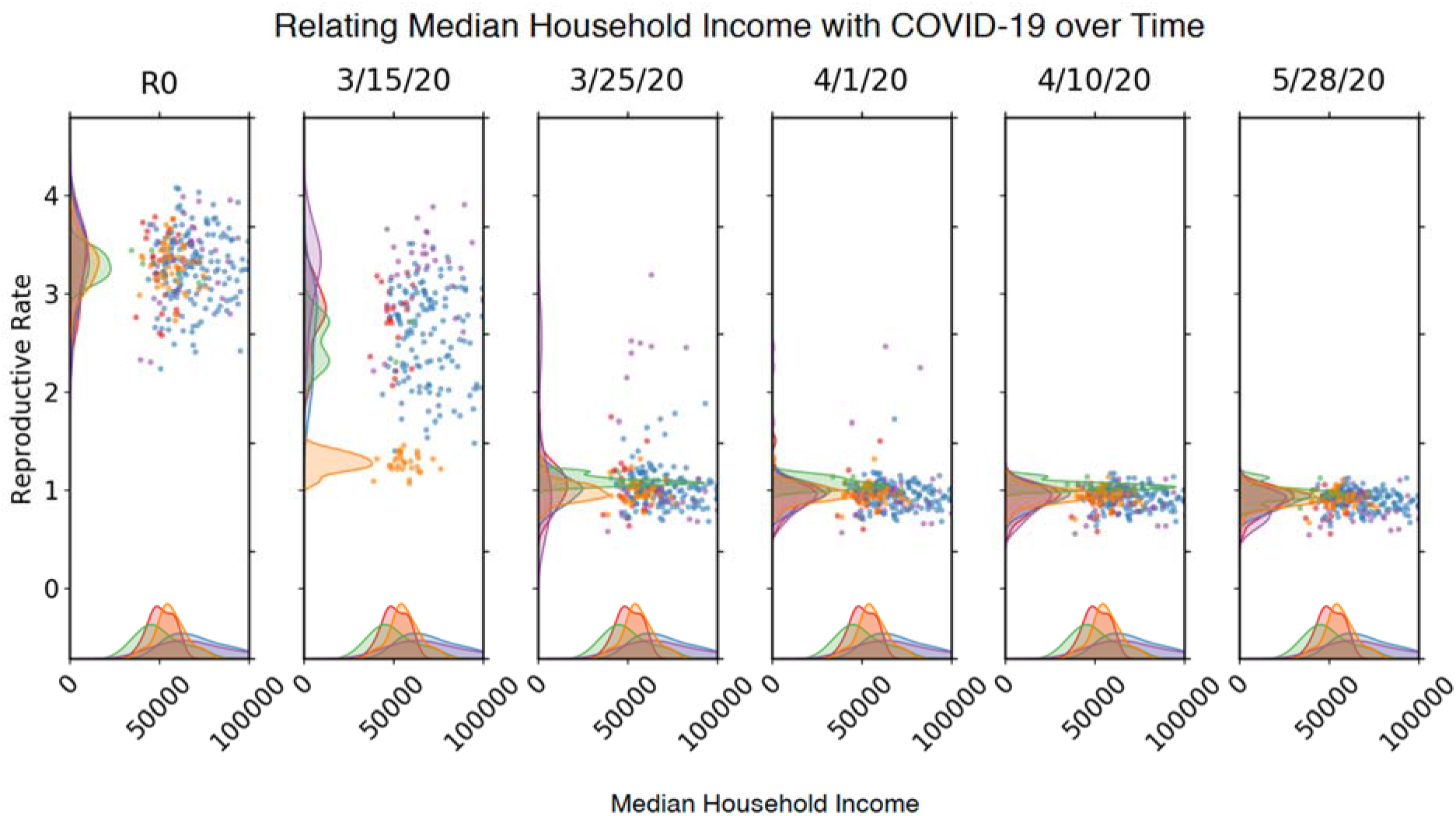
Scatterplot and density distribution plot for counties and super-counties comparing *R_t_* over time to median household income. Colors indicate which cluster the county or super-county belongs in, as indicated by Figure 4.

**Figure 6:**
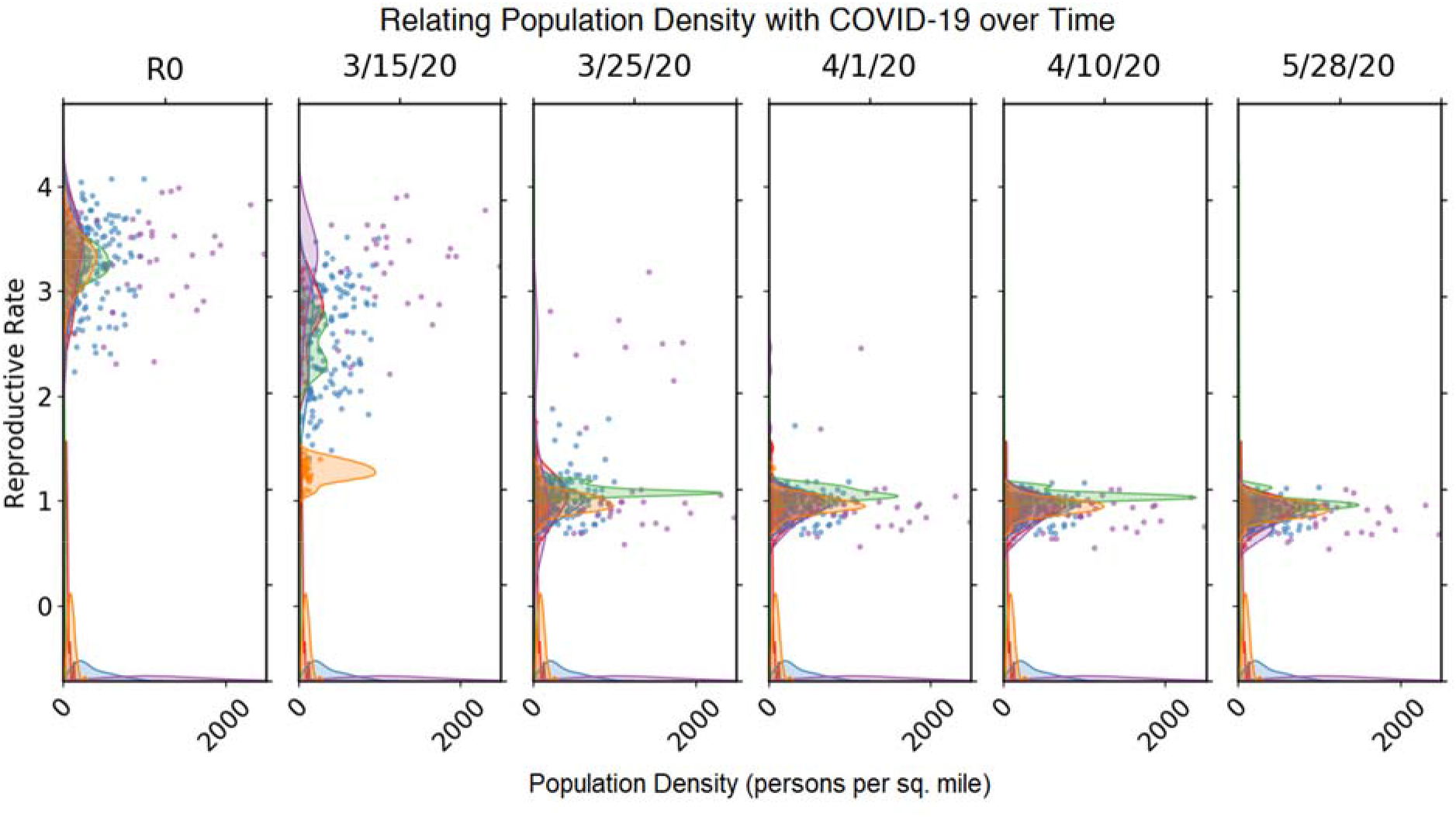
Scatterplot and density distribution plot for counties and super-counties comparing *R_t_* over time to population density. Colors indicate which cluster the county or super-county belongs in, as indicated by Figure 4.

Notably, we exclude ethnic demographics from the variables considered during clustering. because we assume that no direct relationship exists between race and incidence of COVID-19. Rather, such a relationship would be mediated by socioeconomic factors that are already included for similarity assessment.

### Modelling the Effects of NPIs

#### Data Processing

We use cumulative fatality and infection counts from the JHU CSSE COVID-19 Dashboard, which has been tracking COVID-19 since January.^1^ When fitting our model, we use measured fatality rates, which are generally considered more reliable than confirmed infections because of limited testing and the prevalence of asymptomatic cases. Thus, we use population-weighted fatality rates to estimate the true cases count. Obtaining a reasonable estimate for this ratio is crucial to realistically model the numbers of total infections. However, due to asymptomatic cases, undertesting and biased reporting, this parameter cannot be measured directly, but has to be inferred from observable data.^19–21^ Previous studies all report fatality rates with substantial uncertainty but agree on the fact that fatality for COVID-19 depends strongly on the age of the infected person. Therefore, we adapt the fatality rates per age group presented in Verity et al.^19^ for each county with respect to its demographic age distribution. Based on U.S. Census data, a per-county weighted fatality rate is computed using the share of each age group in the overall population.

#### Model

We estimate the effective reproductive rate using a semi-mechanistic Bayesian mechanistic model proposed in Flaxman et al.^5,22^, that infers the impact of a predefined set of interventions and estimates the number of infections over time. The model estimates a county-specific initial reproductive rate *R*_0, *m*_ for the *m*-th county and intervention weights *α_i_*, the effect of which is assumed constant for all counties included in joint optimization. These effects are assumed multiplicative, modeled as

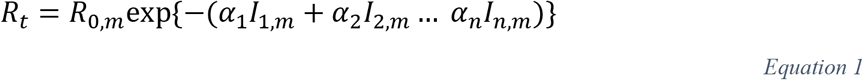

where *m* is the county index and 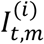 is a binary indicator for intervention *i* being in place at time *t*. The interventions we take into account here are summarized in Table 1.

The model assumes a normal distribution as the prior for the *R*_0_. We set the prior on *R*_0_∽*N*(3.28, ∣*κ*∣) where *κ* ∽ *N*^+^(0, 0.5), The value of 3.28 is in accordance with the analysis presented in Liu et al.^23^

Starting from the time-varying *R_t_*, a latent function of daily infections is modeled depending on a number of factors: a generation distribution *g* with density *g(τ)* that models the time between spread of infection from an individual to the next (approximated as the serial interval distribution), the number of susceptible individuals left in the population, an infection-to-death distribution, and the county-specific time-varying reproduction number that models the average number of secondary infections at any given time. The parameters for the distributions are chosen in accordance to Flaxman et al.^5^

Model fitting is driven by the timeseries of observed daily deaths. These are linked to the modelled number of infections by the county specific weighted fatality rate. The sum of past infections, along with the weighted probability of death gives the number of deaths on a day for a given county. To ensure that the deaths accounted for are from locally acquired infections, we include observed deaths in a county only after the cumulative count has exceeded 10. The seeding of new infections is assumed to be a month prior to that.

All parameters are estimated jointly using an adaptive Hamiltonian Monte Carlo (HMC) sampler in the probabilistic programming language Stan.^24^

### Validation

We propose three validation schemes to validate our approach. First, we show that our model is stable, so a small change in the input produces a small change in the output. Second, we separate our counties and super-counties into train and test sets. We fit models for each cluster as well as for all counties and super-counties on either, both the train and test set or with only the train set. We evaluate using the parameters of the “correct” cluster model to predict fatalities for the held-out regions. Comparing these predictions to those made when the regions are included during training confirms that the model is not over-reliant on each data point. Third, we evaluate how well our model discerns the effects of individual NPIs and discuss biases it may have to attribute more weight to certain NPIs.

#### Validating the Stability of the Model

To show stability in our model, we train the model, withholding three non-consecutive days chosen at random, and compare the predictions for these days to the baseline model fit on full data. Only days with non-zero death counts are selected as potential leave-out candidates. We set the threshold for data selection at counties with 50 cumulative deaths by May 28^th^, which results in 211 counties. (For model validation, we do not use super-counties.) We fit the model over 300 total iterations, with 150 of those as warmup iterations. From the uncertainty distributions obtained as results from the models, we use the expected values of the deaths of the held-out days to compare the two models and report our observations in Figure 7a. The clustering of points around the optimal fit visually confirms that the expected values of the baseline model fit those from the model with held-out days. In Figure 7b, we compare the average county-wise expected value of deaths of the withheld days and observe a more concentrated distribution around the line of optimal fit.

Using the Pearson-correlation coefficient to quantify the similarity between the expected values, we find r-values greater than 0.99 for the held-out days and the average held-out days which suggest that the model is robust against small input perturbations.

**Figure 7:**
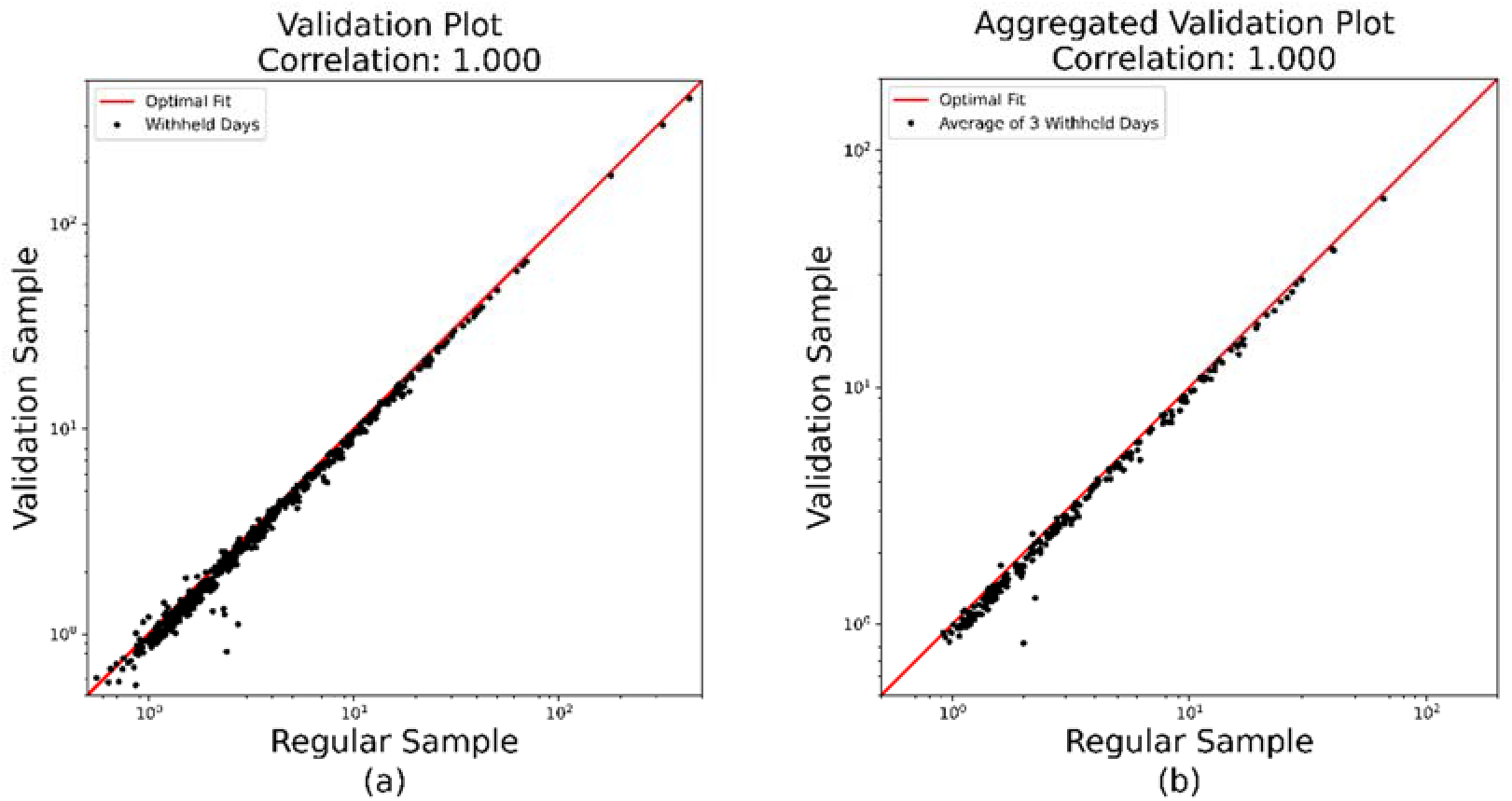
The predicted values of the validation and the baseline model. **(a)** Predictions for withheld days, as shown by the optimal fit, are remarkably close to the true values. **(b)** The three held-out days per county are averaged and compared.

#### Validating the Advantages of Clustering

We show the advantage of cluster-specialized models for quantifying the effects of NPIs by comparing their performance with models trained on different clusters or on the national level. While a clustering of U.S. counties may or may not be interesting, its value related to COVID-19 comes from its ability to identify epidemiologically meaningful differences among various regions in the country. When fitting each model, we withhold a validation region from each cluster and use the remaining counties to fit Equation(1), for both cluster-specialized and baseline models. We then use the learned weights *α_i_* to initialize a fixed-*α* BHI model for each withheld region.

With this scheme, we show the advantage of clustering U.S. counties in three ways. First, we show that within the same cluster, we obtain comparable predictions for a county whether or not it is included in training. Second, we observe that *α* values learned from a different cluster produce substantially different predictions. Finally, we apply the *α* values learned at the national level to similar effect. This demonstrates how cluster-specialized models can reveal trends in the spread of COVID-19 that are not apparent under the assumption that NPIs have universal effect at the national or state level.

Figure 8 shows the first and second validation strategy for cluster 3, which consists of less-populous counties with large land area. For such regions, which necessarily have fewer cases to support local models, it is vital to gain accurate insight from the entire training pool and verify these insights through the aforementioned validation process. Thus, in this illustrative example, we use data up to May 18^th^, at which point the continuing spread of cases was not immediately clear. Indeed, *Figure 8a* shows the predictions obtained for seven counties in Arizona (treated as a single super-county). One can readily observe the model takes into account random outliers while following a general upward trend. Figure 8b exhibits less awareness of outliers but still follows the same trend, despite using α values obtained from training on cluster 3 with these same counties withheld. On the other hand, Figure 8c shows the predictions made when using the wrong α values, from cluster 4, which consists of densely populated city centers. As can be seen, this results in a markedly different prediction for the trend of the disease than actually occurred up to May 28^th^ in Figure 8b. This is a critical difference, which could result in very different decisions for implementing or removing NPIs.

**Figure 8:**
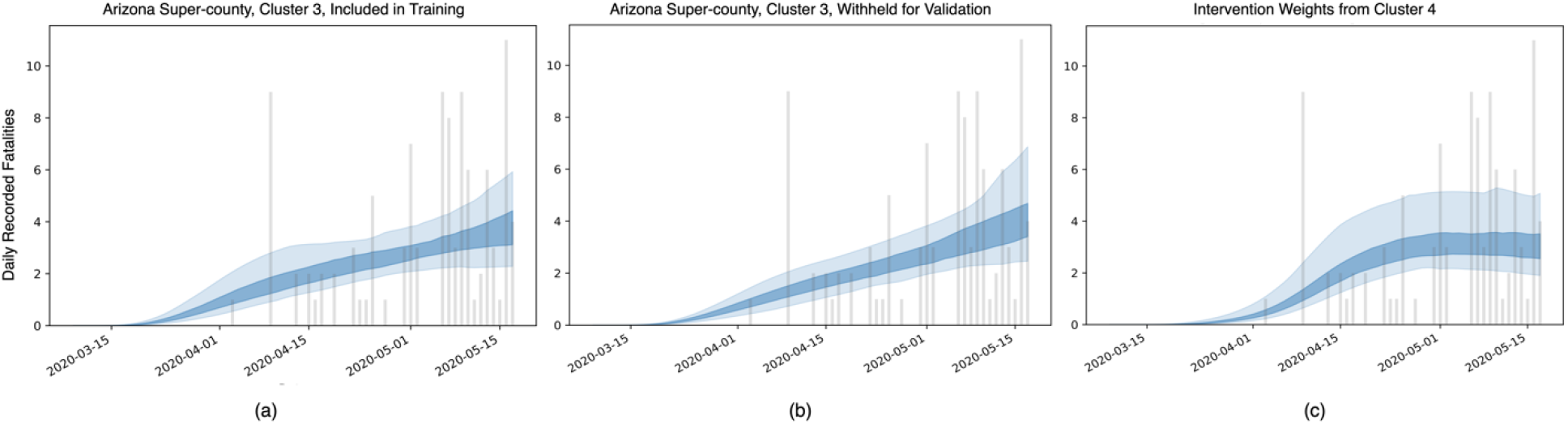
The fatalities predicted by our model for seven counties in Arizona (treated as a single super-county), with fatalities data from Apache, Cochise, La Paz, Mohave, Navajo, Yavapai, and Yuma counties, using data up to May 18^th^. **(a)** When the super-county is included in training, the cluster-specialized BHI model incorporates outlier events in deaths from COVID-19 while still describing the overall trend. **(b)** When the super-county is withheld from training but uses *α* values from the same cluster, the model still captures the same overall trend of increasing deaths. **(c)** On the other hand, using values from a different cluster makes it appear as though the curve has flattened, even though **(d)** shows the continued upward trend based on data up to May 28^th^.

Finally, we compare the performance of our cluster-specialized model with the baseline model, which uses all eligible counties for training. In many cases, this difference is slight, and both the baseline and cluster-specialized models exhibit similar trends for a given region. However, for certain areas the difference can be profound. Take, for example, the District of Columbia, which has become a significant hot-spot for COVID-19. The baseline model, shown in *Figure 9a*, has only recently flattened, whereas the cluster-specialized model in *Figure 9b* reflects the reality that efforts to combat the disease have had much greater effect, significantly reducing the number of deaths. This is because when forced to accommodate counties across the U.S., the baseline model emphasizes federal guidelines with a mean α value of 0.804 (see *Table 3*). On the other hand, our cluster-specialized model found that stay-at-home orders were much more effective for counties in cluster 4, with a mean α value of 0.963. This difference illustrates the advantage of specializing a BHI model based county-level characteristics; it allows the model to include a greater number of counties within separate clusters while not being forced to overgeneralize and, in doing so, compromise some certainty.

**Figure 9:**
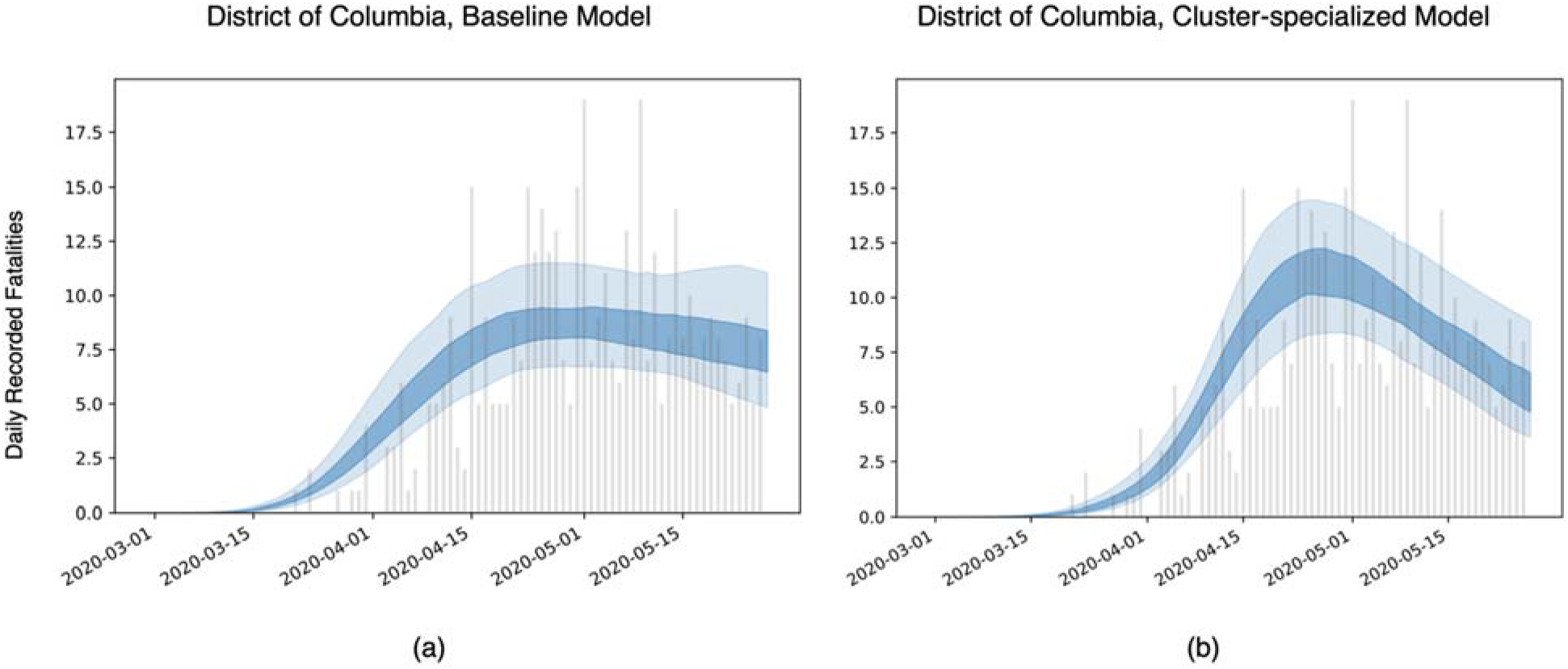
Predicted fatalities in Washington D.C. up to May 28^th^, 2020. **(a)** The fatalities predicted by the baseline model, which observes all eligible counties during training. **(b)** The fatalities predicted by a cluster-specialized model, showing a very different trend in the course of the spread of COVID-19. Although this difference is more pronounced for the District of Columbia than for most regions, it underscores the potential for misleading predictions when the United States’ heterogeneity is not taken into account.

### Disentanglement

One drawback of thes model is that it cannot disentangle implementations that came into effect at the same time. For example, states often closed public schools at the same time as federal guidelines were issued so it is difficult to discern the individual effect on reducing *R_t_*. Additionally, in *Table 5*, we observe that banning gatherings of 50 or more people often occurs at the same time as banning gatherings 500 or more, and restaurants and entertainment venues are often closed together. This suggests that these pairs of interventions may not or be poorly disentangled.

**Table 5:**
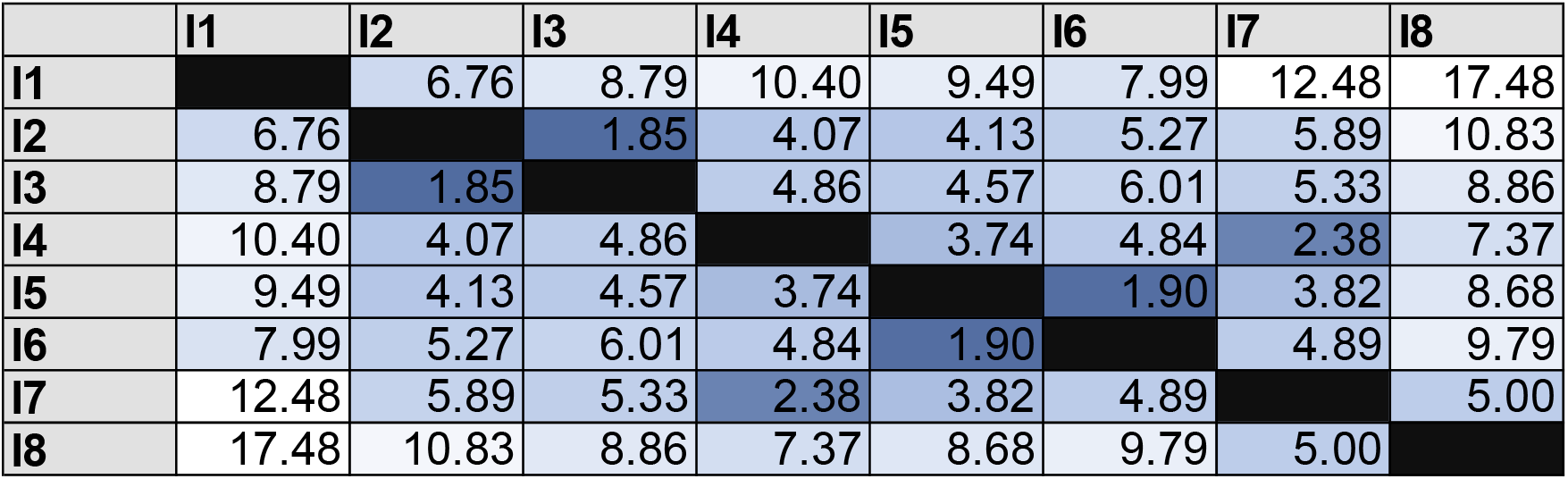
How far apart on average each intervention is implemented from each other. The interventions are in the order defined in Table 2.

To further investigate the model’s ability to disentangle intervention weights, we create simulated trajectories of counties’ deaths and cases counts based on their *R_0_* and the dates on which the interventions came into effect. Using all counties that have more than 50 cumulative deaths on May 28^th^ without super-counties, we seed each county with 200 cases in each of the first 6 days. To simulate county-specific trajectories, we construct a sets of generated timeseries. In the first set, we assign intervention weights *α_i_* to be randomly generated from a Gamma distribution, the same distribution as our prior on the Bayesian mechanistic model adjusted to be in the range of our learned weights. We then calculate what the *R_t_* on each day must have been based on the *R*_0_ and the interventions in place. Once we have the seeded infection and the *R_t_* trajectory for each county, we can calculate daily infections and thus expected fatalities. Using the simulated trajectories, we fit the model. *Table 6* compares the weights used for generation with the weights that the model learned.

**Table 6:** By setting the intervention weights, we can generate simulated timeseries of cases and deaths counts and have the model learn the weights. The learned values differ from the ground truth intervention weights, showing that the model does not disentangle the contribution of each intervention well in cases where interventions were implemented close together..

|  | Intervention weights | Learned weights |
| --- | --- | --- |
| I1 | 0.230 | 0.696 (0.679) |
| I2 | 0.093 | 0.051 (0.068) |
| I3 | 0.128 | 0.122 (0.101) |
| I4 | 0.029 | 0.087 (0.131) |
| I5 | 0.007 | 0.103 (0.179) |
| I6 | 0.321 | 0.271 (0.240) |
| I7 | 0.558 | 0.165 (0.240) |
| I8 | 0.011 | 0.030 (0.070) |

We observe that the effects of individual NPIs are not well disentangled in general. The model tends to attribute more weight to few NPIs rather than spread out the weight evenly. Specifically, the model tends to put more weight on stay-at-home orders. This may be because interventions I2-I8 are often implemented close together (see *Table 5*) and it is difficult to attribute effect to any single one of them on a national scale. While we can conclude that the trajectories the model predicts are reliable, due to their match to measured death, and therefore the overall decrease in *R_t_* is reliable, attributing decreases to any individual NPI is challenging whenever the difference in implementation date is small. This observation seems to be in line with the similarly large confidence intervals reported in previous work on varied models and regions.^2–7^

## Data Availability

All data used in the manuscript is available from public sources.

https://github.com/JieYingWu/COVID-19_US_County-level_Summaries

https://coronavirus.jhu.edu

## Authors’ Contributions

Jie Ying Wu worked on the modeling, disentanglement, and coordinating efforts. Benjamin D Killeen worked on clustering and validating the epidemiological advantage of clustering. Philipp Nikutta worked on processing the data and validating the stability of the model. Mareike Thies did the background research and calculated the weighted fatalities. Anna Zapaishchykova created the methods overview diagram as well as the plots over the features. Shreya Chakraborty worked on processing the data and disentanglement. Mathias Unberath developed the initial concept and direction for this work. Jie Ying Wu, Benjamin D Killeen, and Mathias Unberath wrote the manuscript.

## Acknowledgements

This article was supported in part by institutional funds provided by the Malone Center for
Engineering in Healthcare.

## References

1. Dong, E., Du, H. & Gardner, L. An Interactive Web‐Based Dashboard to Track COVID‐19 in Real Time. Lancet Infect. Dis. 20, 533–534 (2020).

2. Lai, S. et al. Effect of non‐pharmaceutical interventions for containing the COVID‐19 outbreak in China. medRxiv (2020) doi:10.1101/2020.03.03.20029843.

3. Davies, N. G. et al. The effect of non‐pharmaceutical interventions on COVID‐19 cases, deaths and demand for hospital services in the UK: a modelling study. medRxiv (2020) doi:10.1101/2020.04.01.20049908.

4. Mellan, T. A. et al. Report 21: Estimating COVID‐19 cases and reproduction number in Brazil. medRxiv (2020) doi:10.1101/2020.05.09.20096701.

5. Flaxman, S. et al. Estimating the Number of Infections and the Impact of Non‐Pharmaceutical Interventions on COVID‐19 in European Countries: Technical Description Update. arXiv:2004.11342 [stat] (2020).

6. Vollmer, M. A. C. et al. A sub‐national analysis of the rate of transmission of COVID‐19 in Italy. medRxiv (2020) doi:10.1101/2020.05.05.20089359.

7. Fernández‐Villaverde, J. & Jones, C. I. Estimating and Simulating a SIRD Model of COVID‐ 19 for Many Countries, States, and Cities. (2020).

8. Randolph, H. E. & Barreiro, L. B. Herd Immunity: Understanding COVID‐19. Immunity 52, 737–741 (2020).

9. Kwok, K. O., Lai, F., Wei, W. I., Wong, S. Y. S. & Tang, J. W. T. Herd immunity ‐ estimating the level required to halt the COVID‐19 epidemics in affected countries. J. Infect. 80, e32–e33 (2020).

10. Reynolds, D. Gaussian Mixture Models. in Encyclopedia of Biometrics (eds. Li, S. Z. & Jain, A.) 659–663 (Springer US, 2009). doi:10.1007/978-0-387-73003-5_196.

11. Killeen, B. D. et al. A County‐Level Dataset for Informing the United States’ Response to COVID‐19. arXiv:2004.00756 [physics, q‐bio] (2020).

12. Bialek, S. et al. Severe Outcomes Among Patients with Coronavirus Disease 2019 (COVID‐19) — United States, February 12–March 16, 2020. MMWR. Morb. Mortal. Wkly. Rep. 69, 343–346 (2020).

13. Remuzzi, A. & Remuzzi, G. COVID‐19 and Italy: What Next? Lancet 395, 1225–1228 (2020).

14. Epidemiology Working Group for NCIP Epidemic Response & Chinese Center for Disease Control and Prevention. The epidemiological characteristics of an outbreak of 2019 novel coronavirus diseases (COVID‐19) in China. Zhonghua Liu Xing Bing Xue Za Zhi 41, 145–151 (2020).

15. Lee, P.-I., Hu, Y.-L., Chen, P.-Y., Huang, Y.-C. & Hsueh, P.-R. Are Children Less Susceptible to COVID‐19? J. Microbiol. Immunol. Infect. (2020) doi:10.1016/j.jmii.2020.02.011.

16. Ruan, Q., Yang, K., Wang, W., Jiang, L. & Song, J. Clinical Predictors of Mortality Due to COVID‐19 Based on an Analysis of Data of 150 Patients from Wuhan, China. Intensive Care Med. 46, 846–848 (2020).

17. von Gaudecker, H.-M., Holler, R., Janys, L., Siflinger, B. & Zimpelmann, C. Labour Supply in the Early Stages of the COVID‐19 Pandemic: Empirical Evidence on Hours, Home Office, and Expectations. IZA Discuss. Pap. No. 13158 (2020).

18. van Dorn, A., Cooney, R. E. & Sabin, M. L. COVID‐19 Exacerbating Inequalities in the US. Lancet 395, 1243–1244 (2020).

19. Verity, R. et al. Estimates of the severity of coronavirus disease 2019: a model‐based analysis. Lancet Infect. Dis. 20, 669–677 (2020).

20. Russell, T. W. et al. Estimating the infection and case fatality ratio for coronavirus disease (COVID‐19) using age‐adjusted data from the outbreak on the Diamond Princess cruise ship, February 2020. Euro Surveill. 25, (2020).

21. Rinaldi, G. & Paradisi, M. An empirical estimate of the infection fatality rate of COVID‐19 from the first Italian outbreak. medRxiv (2020) doi:10.1101/2020.04.18.20070912.

22. Flaxman, S. et al. Report 13: Estimating the number of infections and the impact of non‐pharmaceutical interventions on COVID‐19 in 11 European countries. (2020) doi:10.25561/77731.

23. Liu, Y., Gayle, A. A., Wilder‐Smith, A. & Rocklöv, J. The reproductive number of COVID‐19 is higher compared to SARS coronavirus. J. Travel Med. 27, (2020).

24. Carpenter, B. et al. Stan : A Probabilistic Programming Language. J. Stat. Softw. 76, (2017).

